# Causal links among amyloid, tau, and neurodegeneration

**DOI:** 10.1101/2021.07.01.21259866

**Authors:** Murat Bilgel, Dean F. Wong, Abhay R. Moghekar, Luigi Ferrucci, Susan M. Resnick, the Alzheimer’s Disease Neuroimaging Initiative

## Abstract

Amyloid-*β* pathology is associated with greater tau pathology and facilitates tau propagation from the medial temporal lobe to the neocortex, where tau is closely associated with local neurodegeneration. The degree of the involvement of amyloid-*β* versus existing tau pathology in tau propagation and neurodegeneration has not been fully elucidated in human studies. Careful quantification of these effects can inform the development and timing of therapeutic interventions.

We conducted causal mediation analyses to investigate the relative contributions of amyloid-*β* and existing tau to tau propagation and neurodegeneration in two longitudinal studies of individuals without dementia: the Baltimore Longitudinal Study of Aging (*N* = 103, age range 57–96) and the Alzheimer’s Disease Neuroimaging Initiative (*N* = 122, age range 56–92). As proxies of neurodegeneration, we investigated cerebral blood flow, glucose metabolism, and regional volume.

We first confirmed that amyloid-*β* moderates the association between tau in the entorhinal cortex and in the inferior temporal gyrus, a neocortical region exhibiting early tau pathology (amyloid group × entorhinal tau interaction term *β* = 0.488, standard error [SE] = 0.126, *P* < 0.001 in the Baltimore Longitudinal Study of Aging; *β* = 0.619, SE = 0.145, *P* < 0.001 in the Alzheimer’s Disease Neuroimaging Initiative). In causal mediation analyses accounting for this facilitating effect of amyloid, amyloid positivity had a statistically significant direct effect on inferior temporal tau as well as an indirect effect via entorhinal tau (average direct effect = 0.47, *P* < 0.001 and average causal mediation effect = 0.44, *P* = 0.0028 in Baltimore Longitudinal Study of Aging; average direct effect = 0.43, *P* = 0.004 and average causal mediation effect = 0.267, *P* = 0.0088 in Alzheimer’s Disease Neuroimaging Initiative). Entorhinal tau mediated up to 48% of the total effect of amyloid on inferior temporal tau. Higher inferior temporal tau was associated with lower colocalized cerebral blood flow, glucose metabolism, and regional volume, whereas amyloid had only an indirect effect on these measures via tau, implying tau as the primary driver of neurodegeneration (amyloid–cerebral blood flow average causal mediation effect = −0.28, *P* = 0.021 in Baltimore Longitudinal Study of Aging; amyloid–volume average causal mediation effect = −0.24, *P* < 0.001 in Alzheimer’s Disease Neuroimaging Initiative).

Our findings suggest targeting amyloid or medial temporal lobe tau might slow down neocortical spread of tau and subsequent neurodegeneration, but a combination therapy may yield better outcomes.

**Abbreviated summary:** Bilgel et al. report that amyloid pathology is a direct driver of tau spread from the medial temporal lobe to the neocortex and that its contribution is comparable to that of existing tau. Their results suggest that therapies targeting both amyloid and tau early might better prevent downstream neurodegeneration.

## Introduction

Amyloid-*β* (A*β*) and hyperphosphorylated tau protein aggregates are hallmark neuropathologies of Alzheimer’s disease and are observed among individuals over the age of 70 regardless of cognitive status.^1^ Tau pathology spreads from the medial temporal lobe (MTL) to the neocortex primarily via synaptic connections^2–5^ in a process facilitated by A*β*.^6,7^ Once tau pathology spreads to the neocortex, it becomes closely linked with neurodegeneration. The spatial distribution of cortical tau overlaps that of local atrophy.^8^ Tau pathology is also associated locally with other markers of neurodegeneration, including lower cerebral blood flow (CBF),^9,10^ glucose metabolism,^11,12^ and volume.^13^ These neurodegenerative changes are thought to eventually lead to cognitive symptoms.

Prior literature provides strong evidence for the involvement of both A*β* pathology and MTL tau in propagating tau to the neocortex. However, the extent to which A*β* versus existing MTL tau pathology is involved in propagating tau remains unclear. An important consideration in the investigation of this question is the facilitating effect of A*β* on tau spread, which requires modeling the interaction between A*β* and MTL tau in relation to neocortical tau using mediation analysis to accurately quantify their relative contributions. Similarly, while previous research has highlighted tau, rather than A*β*, as the driver of neurodegeneration^14,15^ and speculated that tau may mediate the effects of A*β* on cortical neurodegeneration,^16^ these studies have not formally assessed this using mediation analysis.

It is important to quantify the degree of involvement of A*β* and tau pathologies in the propagation of tau and neurodegeneration as these findings may inform the development and timing of therapeutic interventions to prevent or delay the onset of clinical symptoms. If tau propagation and neurodegeneration are largely driven by existing tau pathology, then interventions to alleviate A*β* pathology would not be expected to prevent cortical spreading of tau or neurodegenerative changes. On the other hand, if A*β* also has a substantial direct effect in the neocortical propagation of tau or neurodegeneration, successful therapies among individuals who already have some MTL tau (approximately 70% of individuals aged 60–65)^17^ would need to target both neuropathologies simultaneously.

In this study, we first verify that A*β* facilitates tau spread from MTL to neocortex, and then focus on two scientific questions: (i) To what extent is neocortical tau accumulation driven by A*β* versus MTL tau pathology? (ii) To what extent is neuronal activity, metabolism, or regional volume loss driven by A*β* versus local tau pathology? We investigate these questions among individuals without dementia in two distinct longitudinal studies, the Baltimore Longitudinal Study of Aging (BLSA) and the Alzheimer’s Disease Neuroimaging Initiative (ADNI), to gain insight into changes that occur early in the disease process. To verify that A*β* facilitates the spread of tau from the MTL to the neocortex, we investigate whether A*β* moderates the association between tau in the entorhinal cortex (EC) and the inferior temporal gyrus (ITG). We focus specifically on ITG for evaluating neocortical tau because it is an early neocortical region exhibiting tau pathology^18^ and has often been used to measure tau pathology among individuals without dementia. To quantify the involvement of A*β* versus MTL tau in propagating tau, we evaluate whether EC tau mediates the association between A*β* and ITG tau using causal mediation analysis. We investigate the local relationships between A*β* and proxies of neurodegeneration in ITG, including CBF, glucose metabolism, and regional volume, and whether these relationships are mediated by ITG tau. Finally, we use longitudinal data to assess the directional assumptions of our causal models. Our study substantially builds upon prior work by evaluating certain modeling assumptions with the goal of moving beyond examining associations to enable a causal interpretation of our findings.

## Materials and methods

We conducted our analyses separately in BLSA and ADNI (adni.loni.usc.edu). These data sets allowed us to assess our scientific questions in two complementary stages of Alzheimer’s disease prior to the onset of dementia: most BLSA participants were cognitively normal, whereas most ADNI participants included in these analyses had mild cognitive impairment (MCI). The study samples included participants without dementia who had apolipoprotein E (*APOE*) genotyping, at least one ^18^F-flortaucipir (FTP) PET scan, as well as a *T*_1_-weighted MRI and an amyloid PET scan within 3.5 years of their baseline FTP PET scan. The radiotracer used for amyloid PET was ^11^C-Pittsburgh compound B (PiB) in BLSA and ^18^F-florbetapir (FBP) in ADNI. Dynamic amyloid PET acquisition in BLSA allowed for the computation of the relative radiotracer delivery parameter *R*_1_, which is a surrogate measure of CBF.^19^ In ADNI, instead of CBF, we used a measure of glucose metabolism as assessed by ^18^F-fluorodeoxyglucose (FDG) PET. Both CBF and glucose metabolism are correlates of neuronal activity,^20^ and were used as proxies of neurodegeneration in our analyses, along with regional volume measurements based on MRI parcellation. ADNI participants without an FDG PET scan within 3.5 years of their baseline FTP PET or of the corresponding amyloid PET scans were excluded from the study sample. Each PET and MRI measure was converted to a *z*-score for the statistical analyses, but their original values were used for the summary table and scatter plots.

Multiple MRI and PET scanner vendors and models were used in ADNI. For both MRI and PET, the ADNI study uses phantom scans for calibration across scanners,^21,22^ employs a standard protocol to ensure compatible image acquisitions, and makes available standardized analysis sets that meet minimum quality control requirements.^23^ For a detailed description of MRI and PET protocols, see adni.loni.usc.edu.

In BLSA, cognitively normal status was based on either (i) a Clinical Dementia Rating^24^ of zero and three or fewer errors on the Blessed Information-Memory-Concentration Test,^25^ and therefore the participant did not meet criteria for consensus conference; or (ii) the participant met criteria for consensus conference and was determined to be cognitively normal based on thorough review of clinical and neuropsychological data. MCI diagnoses were determined according to Petersen criteria.^26^ At enrollment into the PET neuroimaging substudy of BLSA, all participants were free of CNS disease (dementia, stroke, bipolar illness, epilepsy), severe cardiac disease, severe pulmonary disease, and metastatic cancer. The assessment of cognitive status and enrollment criteria in ADNI are described elsewhere.^27^

BLSA research protocols were conducted in accordance with United States federal policy for the protection of human research subjects contained in Title 45 Part 46 of the Code of Federal Regulations, approved by local institutional review boards (IRB), and all participants gave written informed consent at each visit. The BLSA PET substudy is governed by the IRB of the Johns Hopkins Medical Institutions, and BLSA is overseen by the National Institute of Environmental Health Sciences IRB. The ADNI study was approved by the IRBs of all participating institutions, and all participants gave written informed consent.

### MR Imaging

#### BLSA

Magnetization-prepared rapid gradient echo (MPRAGE) scans were acquired on a 3 T Philips Achieva scanner (repetition time (TR)=6.8 ms, echo time (TE)=3.2 ms, flip angle=8°, image matrix=256×256, 170 slices, voxel size=1×1×1.2 mm). We computed anatomical labels and regional brain volumes using Multi-atlas region Segmentation using Ensembles of registration algorithms and parameters,^28^ and intracranial volume (ICV) was determined from intracranial masks computed using a deep learning approach.^29^ We computed ICV-adjusted residuals for ITG volume.^30^

#### ADNI

We used tabular data provided by the ADNI MRI Core at the University of California San Francisco for the ADNI3 dataset for regional volumes. These were obtained with cross-sectional FreeSurfer 6.0 (https://surfer.nmr.mgh.harvard.edu/) processing of 3 T MPRAGE scans based on the Desikan-Killiany atlas.^31^ We computed ICV-adjusted residuals for ITG volume as described for BLSA.

### PET Imaging

#### BLSA

Individually-fitted thermoplastic masks were used for all PET scans in BLSA to minimize head motion and ensure consistent table positioning at follow-up visits.

FTP PET scans were obtained over 30 min on a Siemens High Resolution Research Tomograph (HRRT) scanner starting 75 min after an intravenous bolus injection of approximately 370 MBq of radiotracer. Dynamic images were reconstructed using ordered subset expectation-maximization to yield six time frames of 5 min each with approximately 2.5 mm full width at half maximum (FWHM) at the center of the field of view (image matrix=256×256, 207 slices, voxel size=1.22 mm isotropic). Following time frame alignment, the 80 min to 100 min average PET image was partial volume corrected using the region-based voxelwise method.^32^ The corrected image was then used to compute standardized uptake value ratio (SUVR) images using the inferior cerebellar gray matter as the reference region. Inferior rather than whole cerebellar gray matter was used as the reference given the signal spill over from the occipital cortex to the superior cerebellum.^33^ FTP PET image analysis workflow is described in more detail in Ziontz et al.^34^ We computed the average bilateral SUVR in the EC and ITG.

PiB PET scans were obtained over 70 min on either a GE Advance (181 scans) or a Siemens HRRT (96 scans) scanner immediately following an intravenous bolus injection of approximately 555 MBq of radiotracer. Scans acquired on the GE Advance were reconstructed using filtered backprojection with a ramp filter to yield 33 time frames with approximately 4.5 mm FWHM at the center of the field of view (image matrix=128×128, 35 slices, voxel size=2×2×4.25 mm). Scans acquired on the Siemens HRRT were reconstructed using ordered subset expectation-maximization to yield 33 time frames with approximately 2.5 mm FWHM at the center of the field of view (image matrix=256×256, 207 slices, voxel size=1.22 mm isotropic). Since most PiB PET scans were acquired on the GE Advance, the reconstructed HRRT scans were smoothed with a 3 mm FWHM isotropic Gaussian kernel to bring their spatial resolution closer to that of the GE Advance scans, and then resampled to match the voxel size of the GE Advance PiB PET scans. Following time frame alignment and co-registration with MRI, distribution volume ratio (DVR) and relative radiotracer delivery (*R*_1_) images were computed using a spatially constrained simplified reference tissue model (SRTM) with cerebellar gray matter as the reference region.^35^ Cerebellar gray matter was used as the reference region given that it yields SRTM-based PiB DVRs with low test-retest variability and the highest effect size for distinguishing amyloid positivity compared to other common amyloid PET reference regions.^36^ Mean cortical amyloid burden was calculated as the average of the DVR values in cingulate, frontal, parietal (including precuneus), lateral temporal, and lateral occipital cortical regions, excluding the sensorimotor strip. Leveraging longitudinal PiB PET data available on both GE Advance and HRRT scanners for 79 BLSA participants, we estimated the parameters of a linear model mapping mean cortical DVR values between the GE Advance and HRRT scanners and applied this mapping to all HRRT values to harmonize them with the GE Advance values. ITG *R*_1_ values were also harmonized between GE Advance and HRRT scanners using the same approach. Individuals were categorized as amyloid –/+ based on a mean cortical DVR threshold of 1.06, which was derived from a Gaussian mixture model fitted to harmonized mean cortical DVR values at baseline. PiB PET image analysis workflow is described in further detail in Bilgel et al.^19^

#### ADNI

We used tabulated values provided by the ADNI PET Core at the University of California Berkeley, where image quality control and preprocessing were performed. Here, we provide a brief description of image acquisition and of the processing performed by the ADNI PET Core. In ADNI3, FTP PET scans were acquired over 30 min starting 75 min after an intravenous bolus injection of approximately 370 MBq of radiotracer. Images were reconstructed to yield six time frames of 5 min each. FTP values were partial volume corrected using the geometric transfer matrix method^37^ and inferior cerebellar gray matter was used as the reference region to compute SUVRs. We computed a bilateral entorhinal cortex and a bilateral ITG volume-weighted average FTP SUVR using the tabular data provided in the ADNI database.

FBP PET scans were acquired over 20 min starting 50 min after an intravenous bolus injection of approximately 370 MBq of radiotracer and reconstructed to yield four time frames of 5 min each. A composite of whole cerebellum, brainstem/pons, and eroded subcortical white matter was used as the reference region to compute SUVR images given that it yields more accurate longitudinal change measurements compared to other reference regions and might also perform better in cross-sectional settings since it addresses issues that might stem from participant positioning within the scanner’s field of view.^38^ A global cortical amyloid index was calculated as the mean of frontal, lateral temporal, lateral parietal, and cingulate gray matter regions, and amyloid status was defined using an cutoff of 0.78. This cutoff was calculated by transforming the established whole cerebellum reference-based cutoff of 1.11, which is the upper bound of the 95% confidence interval of the mean in a group of young controls,^39^ into the composite reference region units via linear regression.

FDG PET scans were acquired over 30 min starting 30 min after an intravenous bolus injection. For FDG, we averaged the left and right temporal lobe components of the meta-region of interest (ROI) reported by Landau et al.,^40^ which falls largely within the ITG (but also includes part of the middle temporal gyrus).

### Statistical analysis

We converted each continuous PET measure to *z*-scores using the mean and standard deviation at the visit corresponding to (or closest to) the baseline FTP PET scan for each participant. *z*-scores were computed separately for BLSA and ADNI. In the rest of the Methods section and in the Results, we refer to EC (or ITG) FTP SUVR *z*-score as EC (or ITG) tau, ITG PiB *R*_1_ *z*-score as ITG CBF, ITG FDG SUVR *z*-score as ITG glucose metabolism, and ITG ICV-adjusted volume *z*-score as ITG volume.

### Causal mediation analysis

We first verified that A*β* facilitates the spread of tau from the MTL to the neocortex. To do this, we confirmed that the association between EC and ITG tau differs by amyloid status by assessing if there is an interaction between amyloid status and MTL tau in relation to ITG tau using a linear regression model adjusted for age at FTP PET, sex, *APOE* ε4 positivity, years of education, and 10-year cardiovascular disease (CVD) risk (see Supplement for details).

We then conducted two sets of causal mediation analyses: the first set of analyses investigated the mediating effect of EC tau on the relationship between amyloid and ITG tau, and the second set investigated the mediating effect of ITG tau on the relationship between amyloid and proxies of ITG neurodegeneration. Both sets of causal mediation analyses involved two linear regression models: one for the mediator and another for the outcome. All linear regression models included the exposure (i.e., amyloid status) as an independent variable. The models for the outcome (i.e., ITG tau in the first set and ITG neurodegeneration in the second set) additionally included the mediator (i.e., EC tau in the first set and ITG tau in the second set) as an independent variable, and the models with ITG tau as the outcome variable also included an interaction between amyloid and EC tau as an explanatory variable.

In causal mediation analysis, it is necessary to adjust for covariates that may be confounders of the relationships among the exposure, mediator, and outcome.^41^ Since *APOE* ε4 positivity has been shown to impact both A*β* and MTL tau pathology,^7,42^ we included it as a covariate in all linear regression models. We also included age, sex, education, and CVD risk as covariates given their potential effects, possibly by modifying resistance and resilience to AD, on at least two of the three variables included in our mediation analyses.^43^

Because the outcome model for the first set of analyses included an interaction between the exposure (amyloid) and the mediator (EC tau), the causal mediation analysis involved the estimation of two mediation effects and two direct effects (one per amyloid group). These effects are defined and interpreted within the counterfactual framework.^41^ The average causal mediation effect (ACME; also known as the natural indirect effect) for the amyloid negative (A–) group, denoted ACME_A–_, is the expected difference in the outcome while amyloid status is fixed at A– and the mediator (tau) changes from the value attained or that would have been attained under amyloid negativity to the value under amyloid positivity for each individual. ACME_A+_ is defined similarly, while fixing amyloid status at A+ and changing the mediator from the value attained or would have been attained under A– to the value under A+ for each individual. The average direct effect (ADE; also known as the natural direct effect) for the A– group, denoted ADE_A–_, is the expected difference in the outcome as a result of changing amyloid status from A– to A+ while keeping the mediator at the value observed or that would have been observed under A– for each individual. ADE_A+_ is defined similarly, while keeping the mediator at the value observed or that would have been observed under A+ for each individual. For an A– individual, the value of the mediator had they been A+ is a counterfactual (and vice versa) since it involves imaging a scenario where the mediator is measured under an alternative amyloid status for that individual. ACME and ADE reflect the effect of amyloid on the outcome solely through tau (i.e., in the absence of the outcome’s ability to respond to a change in amyloid status) and not through tau (i.e., in the absence of tau’s ability to respond to a change in amyloid status), respectively. We used non-parametric bootstrap with 5000 Monte Carlo draws to estimate bias-corrected and accelerated confidence intervals for ACME and ADE.

Finally, we assessed the sensitivity of the ACME estimates to the sequential ignorability assumption that there are no unspecified confounders between the mediator and the outcome using a parametric sensitivity analysis based on the residual correlation between the mediator and the outcome.^44^

### Further examination of causal assumptions

In addition to the causal mediation sensitivity analyses described above, we conducted two analyses to further examine the assumptions of our causal modeling. The first of these analyses, based on evaluating conditional independencies between variables, was aimed to assess whether our overarching causal model is inconsistent with the data and if a model with sparser connections between variables is possible. The second analysis was aimed to assess whether longitudinal data provide support for the assumed causal directions among amyloid, tau, and neurodegeneration.

### Conditional independencies

The directed acyclic graph (DAG) motivating our causal mediation analyses is presented in Fig. 1. While there is no statistical approach for proving this graph using observational data, falsification tests can be conducted to determine if the graph is inconsistent with the data by assessing conditional independencies implied by this DAG. The only implied conditional independence by this DAG is that EC tau and ITG neurodegeneration (i.e., CBF in BLSA, glucose metabolism in ADNI, and regional volume) are independent given amyloid, ITG tau, and the covariates (age, sex, *APOE* ε4 positivity, education, and CVD risk). We examined this conditional independency for each neurodegeneration measure using partial correlation.

**Figure 1.**
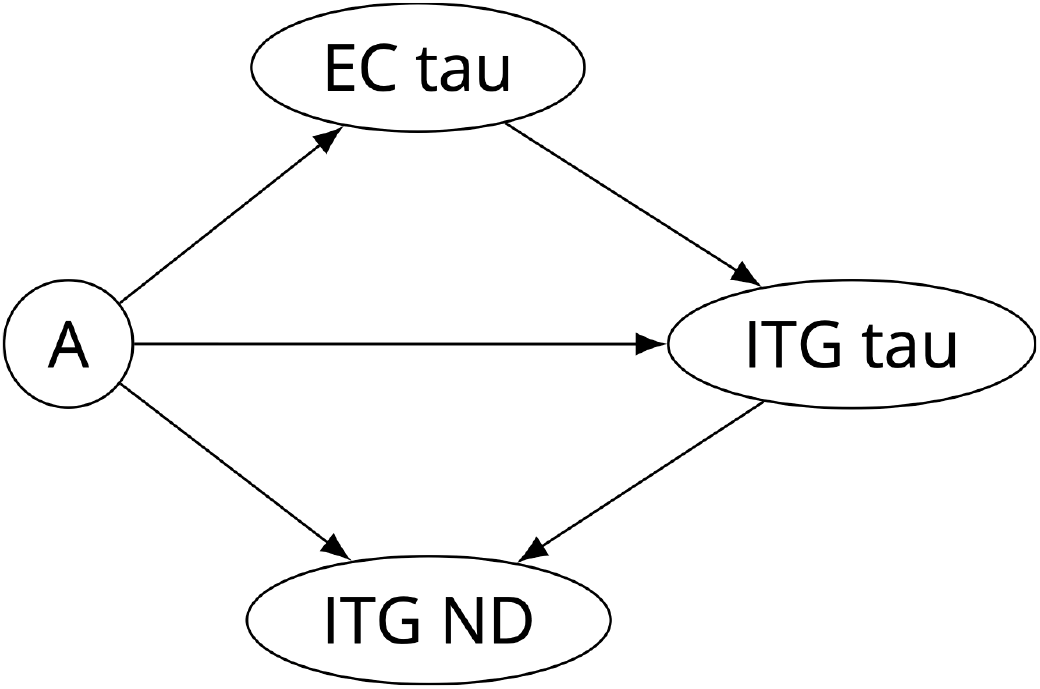
Directed acyclic graph representing the causal model for amyloid (A), tau, and neurodegeneration (ND). As neurodegeneration measures, we investigated cerebral blood flow (in the Baltimore Longitudinal Study of Aging only), glucose metabolism (in the Alzheimer’s Disease Neuroimaging Initiative only), and regional volume. Common covariates for all four nodes (not shown in Figure) include age, sex, *APOE* ε4 positivity, education, and 10-year cardiovascular disease risk. EC = entorhinal cortex, ITG = inferior temporal gyrus.

### Longitudinal analysis

We expected to find evidence using longitudinal data in support of the directions assumed among the variables in our DAG. For example, based on our model assumption that amyloid causally contributes to ITG tau, we expected to find an association between amyloid positivity and the longitudinal rate of tau accumulation in the ITG.

To investigate the rate of change in PET measurements, we assessed visits within ±5 years of baseline FTP PET. Participants did not have dementia at any visit included in longitudinal analyses. In the BLSA sample, there were a total of 167 FTP PET, 277 PiB PET, and 366 MRI scans. In the ADNI sample, there were a total of 211 FTP PET, 194 FDG PET, and 229 MRI scans.

We investigated whether amyloid is associated with the rate of tau accumulation in the EC and ITG using linear mixed effects models. Outcome variables were EC and ITG tau. We included baseline age, time from baseline, sex, years of education, 10-year CVD risk, *APOE* ε4 positivity, amyloid status, and amyloid × time interaction as fixed effects. The model for ITG tau additionally included EC tau and its two-way interactions with amyloid and time as fixed effects. All linear mixed effects models included a random intercept per participant. Random slopes were included only if the total number of observations was greater than the number of random effects to ensure parameter identifiability. The only model that met this criterion and included a random slope per participant was the model for ITG volume in BLSA.

Including race as an additional covariate in BLSA did not affect the statistical significance of any of our cross-sectional or longitudinal findings. We did not investigate race as a covariate in ADNI sample given its limited minority representation.

For all analyses, statistical significance was defined as two-tailed *P* < 0.05. Focusing on a single region (ITG) as the indicator of early neocortical tau pathology allowed us to avoid multiple comparisons. We did not perform multiple comparison correction since we performed one statistical test per family of hypotheses per data set.

### Data availability

Code for performing statistical analyses and generating figures is provided in an open repository (https://gitlab.com/bilgelm/tau_amyloid_neurodegeneration). BLSA data are available upon request from https://www.blsa.nih.gov. All requests are reviewed by the Data Sharing Proposal Review Committee. A deidentified version (with age bands instead of continuous age and excluding *APOE* genotype) of the cross-sectional BLSA data used in the statistical analyses is publicly available (https://doi.org/10.7910/DVN/YFJAZO). ADNI data are available from adni.loni.usc.edu.

## Results

### Participants

Participant characteristics are presented in Table 1 and Supplementary Table 1. 103 BLSA and 122 ADNI participants were included in the analyses. The percentage of participants who were amyloid positive (A+) was 31% in BLSA and 50% in ADNI, which are comparable to the percentages reported in other samples of cognitively normal older individuals and individuals with MCI, respectively.^45^ Regional FTP PET SUVRs were higher in ADNI, but these values are not directly comparable between the studies due to differences in image acquisition and processing.

**Table 1.** Participant characteristics. Continuous values are reported as median (interquartile range) and categorical values are reported as *n* (%). Age, diagnosis, PET measures, and MRI volumes are reported at the visit corresponding to or closest to the baseline FTP PET. Follow-up duration is calculated as the interval between last and first scans included in the longitudinal analyses and its summary statistics are based on participants with at least two scans.

|  | BLSA |  | ADNI |  |
| --- | --- | --- | --- | --- |
|  | A-, *n* = 71 | A+, *n* = 32 | A-, *n* = 61 | A+, *n* = 61 |
| Age at baseline FTP PET (years) | 77 (70, 83) | 78 (73, 87) | 76 (70, 81) | 75 (71, 80) |
| Sex |  |  |  |  |
| Female | 43 (61%) | 16 (50%) | 21 (34%) | 33 (54%) |
| Male | 28 (39%) | 16 (50%) | 40 (66%) | 28 (46%) |
| Race |  |  |  |  |
| Asian or Pacific Islander | 4 (6%) | 1 (3%) | 1 (2%) | — |
| Black | 13 (18%) | 4 (12%) | 2 (3%) | 4 (7%) |
| White | 54 (76%) | 27 (84%) | 57 (93%) | 56 (92%) |
| Other | — | — | 1 (2%) | 1 (2%) |
| *APOE* status |  |  |  |  |
| ε4- | 51 (72%) | 20 (62%) | 52 (85%) | 27 (44%) |
| ε4+ | 20 (28%) | 12 (38%) | 9 (15%) | 34 (56%) |
| Education (years) | 18 (16, 20) | 17 (16, 18) | 18 (16, 19) | 16 (14, 18) |
| 10-year cardiovascular disease risk | 0.16 (0.10, 0.23) | 0.13 (0.08, 0.18) | 0.28 (0.17, 0.37) | 0.17 (0.11, 0.28) |
| Diagnosis at baseline FTP PET |  |  |  |  |
| Cognitively normal | 70 (99%) | 25 (78%) | 11 (18%) | 11 (18%) |
| Mild cognitive impairment | 1 (1%) | 7 (22%) | 50 (82%) | 50 (82%) |
| PET measures at baseline FTP PET |  |  |  |  |
| Entorhinal cortex FTP SUVR (tau) | 1.02 (0.93, 1.1) | 1.07 (1.01, 1.23) | 1.65 (1.5, 2.02) | 2.06 (1.75, 2.58) |
| ITG FTP SUVR (tau) | 1.24 (1.19, 1.32) | 1.37 (1.29, 1.49) | 1.63 (1.57, 1.74) | 1.81 (1.63, 2.21) |
| ITG PiB *R*<sub>1</sub> (cerebral blood flow) | 0.88 (0.86, 0.9) | 0.85 (0.82, 0.9) | — | — |
| ITG FDG SUVR (glucose metab.) | — | — | 1.22 (1.17, 1.31) | 1.21 (1.12, 1.32) |
| ITG volume (ITG-adj. residual, cm<sup>3</sup>) | -0.51 (-1.45, 0.45) | -1.38 (-2.54, 0) | -0.26 (-1.67, 1.1) | -1.61 (-3.12, 0.41) |
| No. FTP PET per participant |  |  |  |  |
| 1 | 41 (58%) | 18 (56%) | 34 (56%) | 26 (43%) |
| 2 | 21 (30%) | 9 (28%) | 18 (30%) | 22 (36%) |
| 3+ | 9 (13%) | 5 (16%) | 9 (15%) | 13 (21%) |
| No. PiB PET per participant |  |  |  |  |
| 1 | 16 (23%) | 8 (25%) | — | — |
| 2 | 21 (30%) | 7 (22%) | — | — |
| 3 | 18 (25%) | 8 (25%) | — | — |
| 4+ | 16 (23%) | 9 (28%) | — | — |
| No. FDG PET per participant |  |  |  |  |
| 1 | — | — | 23 (38%) | 37 (61%) |
| 2 | — | — | 32 (52%) | 20 (33%) |
| 3 | — | — | 6 (10%) | 4 (7%) |
| No. MRI per participant |  |  |  |  |
| 1 | 3 (4%) | 1 (3%) | 15 (25%) | 27 (44%) |
| 2 | 12 (17%) | 8 (25%) | 29 (48%) | 25 (41%) |
| 3 | 26 (37%) | 11 (34%) | 16 (26%) | 9 (15%) |
| 4+ | 30 (42%) | 12 (38%) | 1 (2%) | — |
| FTP PET follow-up duration (years) | 2 (2, 2) | 2 (1, 3) | 2 (1, 2) | 2 (1, 2) |
| PiB PET follow-up duration (years) | 4 (2, 5) | 4 (3, 4) | — | — |
| FDG PET follow-up duration (years) | — | — | 4 (4, 5) | 4 (4, 5) |
| MRI follow-up duration (years) | 4 (4, 6) | 4 (4, 5) | 2 (1, 2) | 1 (1, 2) |

Abbreviations: A<sup>−</sup> = amyloid negative, A<sup>+</sup> = amyloid positive, ADNI = Alzheimer's Disease
Neuroimaging Initiative, *APOE* = apolipoprotein E (gene), BLSA = Baltimore Longitudinal Study of Aging, FDG = <sup>18</sup>F-fluorodeoxyglucose, FTP = <sup>18</sup>F-flortaucipir, ICV-adj. = intracranial volume-adjusted, ITG = inferior temporal gyrus, PiB = Pittsburgh compound B, *R*<sub>1</sub> = relative radiotracer delivery parameter, SUVR = standardized uptake value ratio.

### Causal mediation analyses

#### (1) To what extent is neocortical tau accumulation driven by A*β* versus MTL tau pathology?

As expected based on the visual assessment of the relationship between EC tau and ITG tau by amyloid status (Fig. 2), both amyloid positivity and higher EC tau were associated with higher ITG tau and the A+ group exhibited a stronger correlation between the levels of tau pathology in these two regions (amyloid × EC tau interaction term *β* = 0.488, standard error [SE] = 0.126, *P* < 0.001 in BLSA; *β* = 0.619, SE = 0.145, *P* < 0.001 in ADNI).

**Figure 2.**
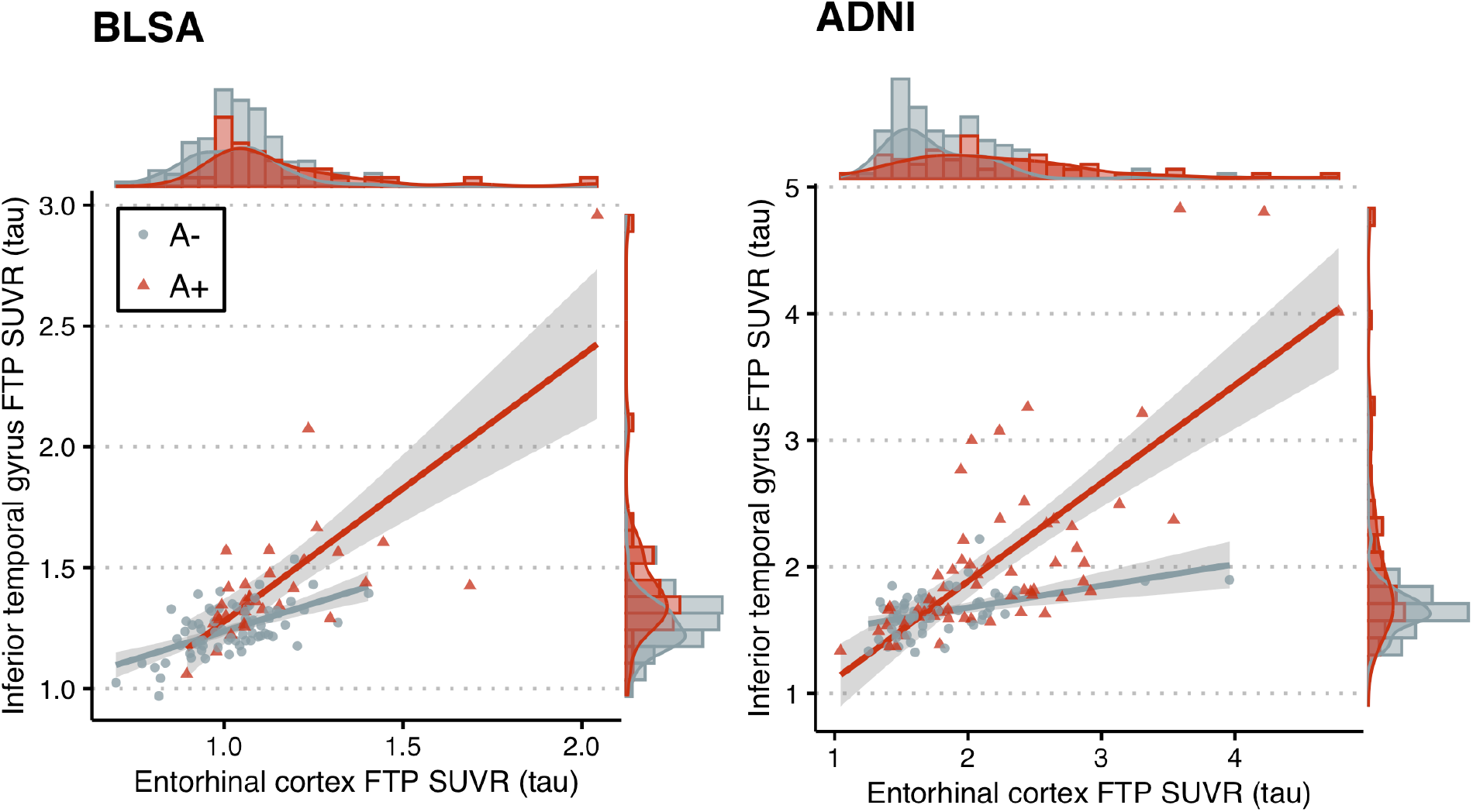
Scatter plots of inferior temporal gyrus vs. entorhinal cortex tau in BLSA and ADNI. In adjusted linear regression models, higher entorhinal tau was associated with higher inferior temporal gyrus tau in both amyloid– (A–) (EC tau effect among A–: *β* = 0.383, standard error [SE] = 0.0972, *t* = 3.94, *P* < 0.001 in BLSA; *β* = 0.260, standard error [SE] = 0.123, *t* = 2.10, *P* = 0.038 in ADNI) and A+ groups (EC tau effect among A+: *β* = 0.871, SE = 0.0818, *t* = 10.6, *P* < 0.001 in BLSA; *β* = 0.879, SE = 0.0766, *t* = 11.5, *P* < 0.001 in ADNI), with the A+ group exhibiting a stronger positive correlation (amyloid × EC tau interaction term *β* = 0.488, SE = 0.126, *t* = 3.86, *P* < 0.001 in BLSA; *β* = 0.619, SE = 0.145, *t* = 4.28, *P* < 0.001 in ADNI). ADNI = Alzheimer’s Disease Neuroimaging Initiative, BLSA = Baltimore Longitudinal Study of Aging, FTP = flortaucipir, SUVR = standardized uptake value ratio.

In causal mediation analyses examining the extent to which neocortical tau accumulation is driven by A*β* versus MTL tau pathology, we found that the effect of amyloid on ITG tau was mediated by EC tau (ACME_A–_ = 0.267, *P* = 0.0028 and ACME_A+_ = 0.607, *P* = 0.0028 in BLSA; ACME_A–_ = 0.122, *P* = 0.0092 and ACME_A+_ = 0.412, *P* = 0.0088 in ADNI) (Fig. 3). In addition to this mediation effect, amyloid had a direct effect on ITG tau (ADE_A–_ = 0.297, *P* = 0.0384 and ADE_A+_ = 0.637, *P* < 0.001 in BLSA; ADE_A+_ = 0.573, *P* < 0.001 in ADNI) (Fig. 3). Estimates are summarized in Supplementary Table 2a. In both datasets, ACME_A+_ was robust to violations of the assumption that there are no unspecified confounders between the mediator (EC tau) and outcome (ITG tau) (Supplementary Fig. 1).

**Figure 3.**
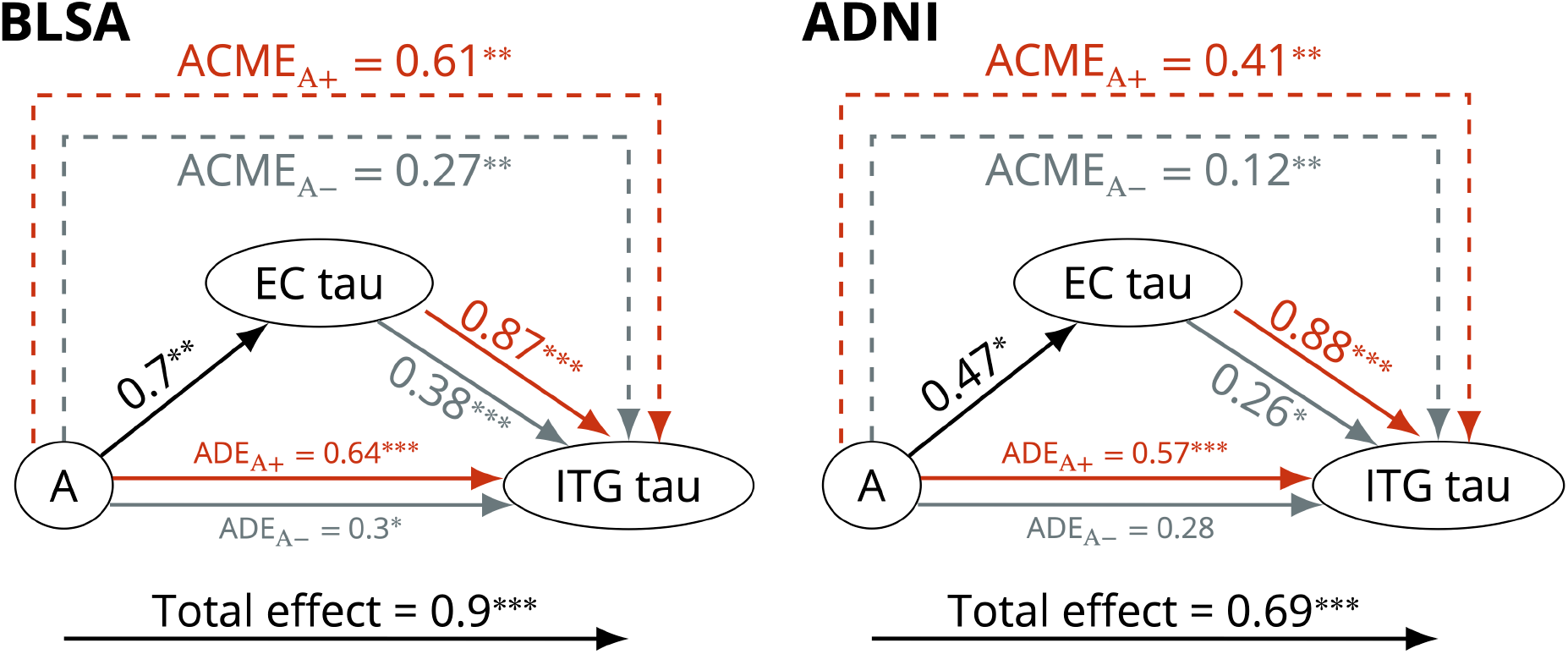
Causal mediation analysis results for the model investigating the relationships among amyloid group (A) and tau in the entorhinal cortex (EC) and the inferior temporal gyrus (ITG) in BLSA (*left*) and ADNI (*right*). The effect of amyloid on ITG tau is mediated by EC tau. Arrow from A to EC tau indicates the linear regression coefficient, and the gray and red arrows from EC tau to ITG tau indicate the linear regression coefficients for the amyloid negative and positive groups, respectively. Both the mediator and the outcome models were adjusted for age, sex, *APOE* ε4 positivity, years of education, and 10-year cardiovascular disease risk. ACME = average causal mediation effect, ADE = average direct effect. *** P *<* .001, ** P *<* .01, * P *<* .05.

#### (2) To what extent are proxies of neurodegeneration driven by A*β* versus local tau pathology?

In separate causal mediation analyses investigating the direct and indirect effects of A*β* on proxies of ITG neurodegeneration, the inclusion of an interaction between the exposure (amyloid) and mediator (ITG tau) did not yield a statistically significant improvement in the residual sum of squares (ANOVA *F* = 2.90, *P* = 0.092 for ITG CBF and *F* = 0.287, *P* = 0.59 for ITG volume in BLSA; *F* = 2.16, *P* = 0.14 for ITG glucose metabolism and *F* = 2.25, *P* = 0.14 for ITG volume in ADNI) for any of the models investigating proxies of ITG neurodegeneration as outcomes, and therefore, this interaction was not included in the final models. Unlike in our first causal mediation analysis, ACME and ADE do not depend on amyloid status in this case because there is no interaction between the exposure and mediator. The effect of amyloid positivity on ITG CBF was mediated via ITG tau in BLSA (ACME = −0.283, *P* = 0.021) (Fig. 4), and the effect of amyloid positivity on ITG volume was mediated via ITG tau in ADNI (ACME = −0.238, *P* < 0.001) (Fig. 4). These ACME estimates were not as robust as our other findings to violations of the assumption of no unspecified confounders between the mediator (ITG tau) and the outcome (ITG CBF or volume) (Supplementary Fig. 2).

**Figure 4.**
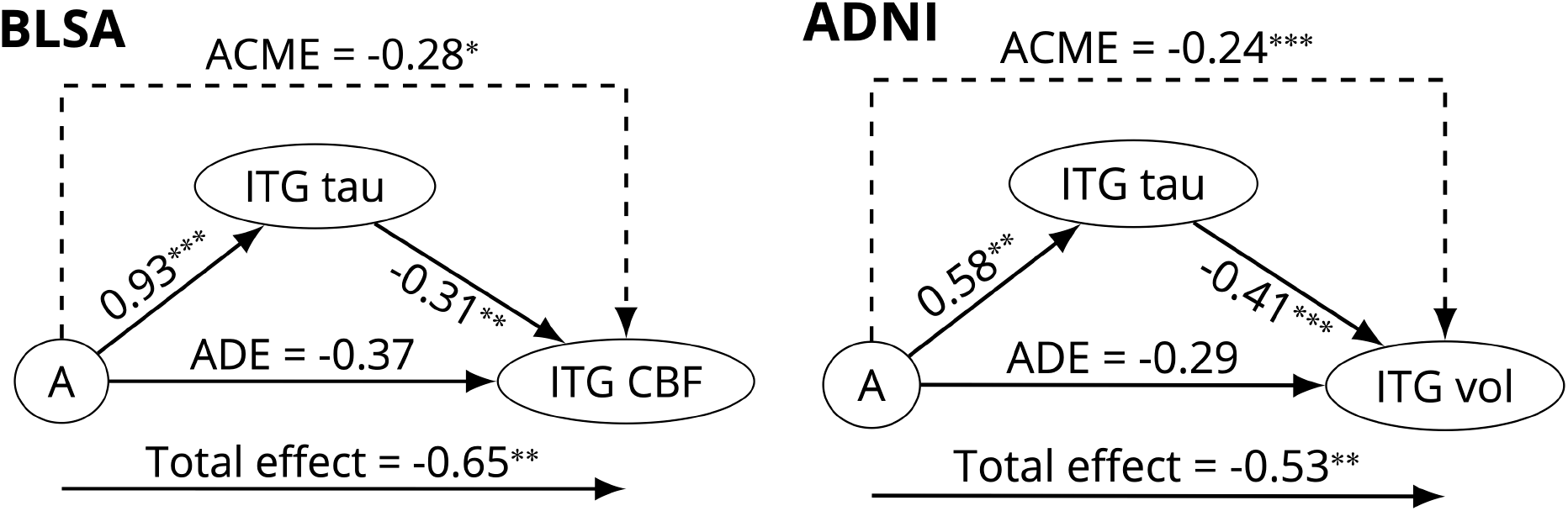
Causal mediation analysis results for the model investigating the relationships among amyloid group (A), tau in the inferior temporal gyrus (ITG), and neurodegeneration in the ITG. The effect of amyloid on ITG cerebral blood flow (CBF) (in BLSA) and on ITG volume (in ADNI) was mediated via ITG tau. Arrows from A to ITG tau and from ITG tau to ITG CBF indicate the linear regression coefficients for these associations. Both the mediator and the outcome models were adjusted for age, sex, *APOE* ε4 positivity, years of education, and 10-year cardiovascular disease risk. ACME = average causal mediation effect, ADE = average direct effect. *** P *<* .001, ** P *<* .01, * P *<* .05.

We do not report the results of the causal mediation or sequential ignorability sensitivity analysis investigating ITG volume in BLSA or ITG glucose metabolism in ADNI since the associations between amyloid and these variables were small and not statistically significant (total effect on ITG volume in BLSA = −0.26, *P* = 0.24; total effect on ITG glucose metabolism in ADNI = −0.059, *P* = 0.76; Supplementary Tables 2b and 2c).

We repeated all causal mediation analyses using continuous amyloid measures and additionally in ADNI, using amyloid group designations based on a whole cerebellar reference region. Results were highly consistent with our main results (Supplementary Tables 3 and 4).

### Further examination of causal assumptions

#### Conditional independencies

The partial correlation between EC tau and ITG CBF given amyloid status and all covariates was small and not statistically significant in BLSA (*ρ* = 0.127, *P* = 0.22). Similarly, the partial correlation between EC tau and ITG glucose metabolism was small and not statistically significant in ADNI (*ρ* = −0.175, *P* = 0.061). The partial correlation between EC tau and ITG volume given amyloid status and all covariates was also small and not statistically significant in both BLSA (*ρ* = 0.161, *P* = 0.12) and ADNI (*ρ* = −0.18, *P* = 0.054). These results agree with the conditional independency implied by our DAG. We also assessed more parsimonious versions of this model (i.e., DAGs with fewer edges) by testing their implied conditional independencies using partial correlations, but none satisfied all implied conditional dependencies (Supplementary Fig. 3). Of note, even though the indirect effect of amyloid on proxies of neurodegeneration was not statistically significant in the causal mediation analyses, removing the arrow from amyloid to neurodegeneration in the DAG yielded a conditional independence (i.e., that amyloid and ITG neurodegeneration are independent given ITG tau and covariates) that was not consistent with the data.

#### Longitudinal analyses

In BLSA, we did not find a statistically significant association between the rate of change in EC tau and amyloid status (*β* = 0.079, SE = 0.0605, *P* = 0.197), or between the rate of change in ITG tau and amyloid status (*β* = 0.0427, SE = 0.0391, *P* = 0.279) or baseline EC tau (*β* = −0.0297, SE = 0.0273, *P* = 0.282). In ADNI, A+ individuals had steeper increases in EC tau (*β* = 0.0806, SE = 0.0324, *P* = 0.015) and ITG tau (*β* = 0.0714, SE = 0.0299, *P* = 0.02). Higher baseline EC tau was associated with a steeper increase in ITG tau over time (*β* = 0.0719, SE = 0.0194, *P* < 0.001).

We also investigated whether ITG tau is associated with the rate of change in ITG CBF, glucose metabolism, or volume. The outcome variable was ITG CBF (in BLSA), glucose metabolism (in ADNI), or volume. We did not find an association between amyloid status and the rate of change in ITG CBF (*β* = 0.0147, SE = 0.0313, *P* = 0.64 in BLSA), glucose metabolism (*β* = −0.0187, SE = 0.0398, *P* = 0.64 in ADNI), or volume (*β* = −0.0129, SE = 0.0228, *P* = 0.572 in BLSA; *β* = 0.00133, SE = 0.0408, *P* = 0.975 in ADNI). Higher baseline ITG tau was associated with a steeper decrease in ITG glucose metabolism (*β* = −0.0706, SE = 0.041, *P* = 0.09) and ITG volume over time (*β* = −0.0487, SE = 0.0139, *P* < 0.001 in BLSA; *β* = −0.0702, SE = 0.0248, *P* = 0.006 in ADNI), but not with the rate of change in ITG CBF (*β* = 0.0291, SE = 0.019, *P* = 0.128).

## Discussion

In both BLSA and ADNI, greater EC tau was associated with greater ITG tau cross-sectionally and this association was stronger among amyloid positive individuals. These findings agree with prior reports that A*β* facilitates the spread of tau from the MTL to the neocortex.

Amyloid’s association with ITG tau was in part mediated by EC tau: EC tau mediated 48% of the total effect in BLSA and 38% in ADNI. This mediation effect was robust to the sequential ignorability assumption of causal mediation analysis, meaning that any unspecified confounder of the mediator and outcome relationship must have substantially correlated effects on these two variables for our reported mediation effects to become statistically non-significant.

Higher ITG tau was associated with several proxies of neurodegeneration in the same region, including CBF, glucose metabolism, and volume (in ADNI only). While we did not find a direct effect of amyloid positivity on neurodegeneration, it had an indirect effect via tau pathology on CBF (BLSA) and volume (ADNI). The indirect effect of amyloid via ITG tau accounted for 44% of the total effect on ITG CBF in BLSA and 45% of the total effect on ITG volume in ADNI, but the statistical significance of these indirect effect estimates was sensitive to the assumption that there were no unspecified confounders of ITG tau and neurodegeneration.

In longitudinal analyses, amyloid positivity was associated with steeper increases in EC tau and greater EC tau at baseline was associated with steeper increases in ITG tau in ADNI. The lack of statistically significant amyloid associations in BLSA might be due to insufficient statistical power to detect small longitudinal effects among cognitively normal individuals.^46^ Even though we controlled for *APOE* ε4 carrier status in our longitudinal analyses, the greater proportion of *APOE* ε4 carriers in ADNI compared to BLSA might also explain the different longitudinal findings in the two datasets. Greater ITG tau at baseline was associated with steeper decreases in glucose metabolism in the same region. The finding that greater ITG tau at baseline is associated with faster volume loss was shared between BLSA and ADNI.

Our findings may have therapeutic implications. The causal mediation results suggest that treating EC tau alone would likely be insufficient in preventing the neocortical spread of tau, since the direct path between amyloid and ITG tau would not be affected. Similarly, preventing amyloid alone among individuals who already have some level of EC tau would also likely be insufficient, since EC tau would continue to spread to the neocortex. However, given the synergistic association between amyloid and EC tau in neocortical propagation, targeting either pathology would be expected to slow down the spread of tau to the neocortex. A recent clinical trial of donanemab, an anti-amyloid monoclonal antibody, conducted among patients with early symptomatic Alzheimer’s disease failed to find a difference in global tau load between treatment and placebo groups, but there was a greater reduction in tau accumulation in frontal and temporal regions.^47^ An amyloid intervention earlier in the disease course, combined with a tau-targeting therapy, might yield more pronounced effects on neocortical tau.

Tau, rather than amyloid, might be the main driver of neurodegeneration. In the recently completed phase 2 trial among patients with mild AD of AADvac1, an active immunotherapy against pathological tau, neurodegeneration was slower in the treatment group, as evidenced by an attenuated increase in neurofilament light.^48^ This clinical trial result supports our model assumption that tau is a contributor but also aligns with previous reports that tau pathology does not explain the entire degree of neurodegeneration.^49^ We did not observe an interaction between amyloid and neocortical tau in relation to neurodegeneration, while such a synergistic relationship has been reported in transgenic mice^6^ and human studies.^12,50^

Our finding that tau pathology mediated the association of amyloid with volume in ADNI but not BLSA might be explained by differences in the composition of the samples: most ADNI participants included in our analyses had MCI, whereas most BLSA participants were cognitively normal. This difference between the two samples suggests that the detrimental effects of tau pathology on volume loss might not be evident until later stages of disease. Supporting this hypothesis, a simulation study based on quantitative neuron loss and neurofibrillary tangle formation data suggested that neurons harboring may survive for about two decades.^51^ Our findings in BLSA might reflect differences in the temporal sequence of changes. CBF might exhibit neuropathology-associated changes while individuals are cognitively normal, prior to declines in regional brain volumes.^52^ This could explain the absence of a cross-sectional association of tau with volume in BLSA and the presence of an association with CBF.

Our longitudinal findings in ADNI provide support for the causal directions assumed in our DAG between amyloid and tau, and agree with the findings of several observational studies where A*β* was associated with faster tau accumulation over time.^53–57^ In analyses adjusted for tau, we did not find an association between amyloid positivity and rate of change in any of the neurodegeneration measures we assessed. The lack of an association suggests that the direct effect of fibrillar amyloid on neurodegenerative change is small and requires larger samples to be detected, or that fibrillar amyloid is not a direct player in CBF, glucose metabolism, or volume loss changes among individuals without dementia.

Limitations of our study include the small samples and the lack of measures for other factors implicated in tau spread (i.e., neuroinflammation or synaptic markers), which prevents the analysis of competing mechanisms. Functional^58^ and structural^59^ connections between regions have been shown to be associated with the propagation of tau and could be important mediators of the association between amyloid and neocortical tau. We used binary amyloid status based on fibrillar A*β* burden, but a measure of soluble A*β* could have been more appropriate given the evidence that soluble forms induce tau hyperphosphorylation^60^ and may be more closely involved in neurodegeneration rather than fibrillar amyloid. However, concentrations of soluble A*β* oligomers correlate with the amount of A*β* plaques, albeit weakly, among individuals without dementia.^61^ Larger samples of A+ individuals will enable a more thorough investigation of the dose effect of amyloid burden. It is also possible that there are mechanisms by which cortical fibrillar amyloid deposition can influence MTL tau pathology; for example, amyloid pathology is associated with axonal dystrophy,^62^ which might be a mechanism by which neocortical amyloid exerts retrograde effects. While we assumed a unidirectional association between amyloid and tau, it is possible that these pathologies exist within a feedback loop. One study suggested that amyloid and tau pathology reinforce each other.^6^ There are differing schools of thought about the sequence of changes in CBF and tau.^63^ It is unclear whether PET measures of CBF would reliably detect the types of early changes that are hypothesized to lead to tau pathology. Extensive longitudinal data will be necessary to determine the direction of these associations. The variables we used to assess neurodegeneration are not direct measures but rather proxies. Cerebral amyloid angiopathy, which can be detected using amyloid PET but with low specificity, has direct effects on cerebrovascular function. This confound might impact the interpretation of cerebral blood flow as an indicator of neuronal activity. Given that neuropathology and neurodegeneration are putative causes of cognitive impairment, we might in essence be conditioning on a collider and creating spurious correlations among amyloid, tau, and neurodegeneration by studying a sample with a high percentage of MCI as in our ADNI sample. As a result, we may have overestimated the causal effects in ADNI. Using cross-sectional rather than longitudinal data in causal inference may have yielded estimates conflated by individual differences.

An important strength of our study is the replication of many of our findings in two longitudinal cohorts. In particular, the causal mediation analysis findings regarding amyloid-facilitated propagation of tau from the MTL to the neocortex were highly consistent in BLSA and ADNI despite differences in sample composition. Based on this consistency, we expect these findings to be generalizable to similar samples of individuals without dementia.

In conclusion, our study confirms the facilitating role of amyloid in the propagation of tau from the MTL to the neocortex among cognitively normal individuals and in MCI. Since entorhinal tau mediated only up to half of the association of amyloid with neocortical tau, we speculate that interventions to reduce only entorhinal tau or only amyloid among individuals who already have MTL tau might slow but not fully prevent the neocortical spread of tau. Preventing neocortical tau early might be an important element in preventing cortical neurodegeneration, including lower CBF, hypometabolism, and volume loss. It will be important to examine the potential impact of reducing amyloid and tau on cognitive decline in future studies and to determine if preventing the spread of tau from the MTL to the neocortex would yield clinical benefit.

## Data Availability

Code for performing statistical analyses and generating figures is provided in an open repository (https://gitlab.com/bilgelm/tau_amyloid_neurodegeneration). BLSA data are available upon request from https://www.blsa.nih.gov. All requests are reviewed by the BLSA Data Sharing Proposal Review Committee. A deidentified version (with age bands instead of continuous age and excluding APOE genotype) of the cross-sectional BLSA data used in the statistical analyses is publicly available (https://doi.org/10.7910/DVN/YFJAZO). ADNI data are available from adni.loni.usc.edu.

http://adni.loni.usc.edu

https://www.blsa.nih.gov

## Abbreviations

A*β*: Amyloid-beta
ACME: Average causal mediation effect
ADE: Average direct effect
ADNI: Alzheimer’s Disease Neuroimaging Initiative
*APOE*: Apolipoprotein E
BLSA: Baltimore Longitudinal Study of Aging
CBF: Cerebral blood flow
DAG: Directed acyclic graph
DVR: Distribution volume ratio
EC: Entorhinal cortex
FBP: ^18^F-florbetapir
FDG: ^18^F-fluorodeoxyglucose
FTP: ^18^F-flortaucipir
FWHM: Full width at half maximum
HRRT: High resolution research tomograph
ICV: Intracranial volume
ITG: Inferior temporal gyrus
MCI: Mild cognitive impairment
MPRAGE: Magnetization prepared rapid gradient echo
MTL: Medial temporal lobe
PiB: ^11^C-Pittsburgh compound B
*R*_1_: Relative radiotracer delivery parameter
SE: Standard error
SUVR: Standardized uptake value ratio
TE: Echo time
TR: Repetition time

## Acknowledgements

We thank Dr. Andrea Shafer and Dr. Fırat Bilgel for their thoughtful feedback. Graphical abstract was created with BioRender.com.

## Funding

This study was supported by the Intramural Research Program of the National Institute on Aging, NIH.

Data collection and sharing for the ADNI sample was funded by the Alzheimer’s Disease Neuroimaging Initiative (ADNI) (National Institutes of Health Grant U01 AG024904) and DOD ADNI (Department of Defense award number W81XWH-12-2-0012). ADNI is funded by the National Institute on Aging, the National Institute of Biomedical Imaging and Bioengineering, and through generous contributions from the following: AbbVie, Alzheimer’s Association; Alzheimer’s Drug Discovery Foundation; Araclon Biotech; BioClinica, Inc.; Biogen; Bristol-Myers Squibb Company; CereSpir, Inc.; Cogstate; Eisai Inc.; Elan Pharmaceuticals, Inc.; Eli Lilly and Company; EuroImmun; F. Hoffmann-La Roche Ltd and its affiliated company Genentech, Inc.; Fujirebio; GE Healthcare; IXICO Ltd.; Janssen Alzheimer Immunotherapy Research & Development, LLC.; Johnson & Johnson Pharmaceutical Research & Development LLC.; Lumosity; Lundbeck; Merck & Co., Inc.; Meso Scale Diagnostics, LLC.; NeuroRx Research; Neurotrack Technologies; Novartis Pharmaceuticals Corporation; Pfizer Inc.; Piramal Imaging; Servier; Takeda Pharmaceutical Company; and Transition Therapeutics. The Canadian Institutes of Health Research is providing funds to support ADNI clinical sites in Canada. Private sector contributions are facilitated by the Foundation for the National Institutes of Health (www.fnih.org). The grantee organization is the Northern California Institute for Research and Education, and the study is coordinated by the Alzheimer’s Therapeutic Research Institute at the University of Southern California. ADNI data are disseminated by the Laboratory for Neuro Imaging at the University of Southern California.

## Competing interests

MB, LF, ARM, and SMR declare that they have no competing interests. DFW is a former President of the Brain Imaging Council of the Society of Nuclear Medicine and Molecular Imaging. DFW receives contract funding from LB Pharmaceuticals and in-kind contributions from Roche Neuroscience, Avid Radiopharmaceuticals/Eli Lilly and Company, and Cerveau Technologies.

## Supplementary material

Supplementary material is available at *Brain* online.

## Appendix 1

A complete listing of ADNI investigators can be found at: http://adni.loni.usc.edu/wp-content/uploads/how_to_apply/ADNI_Acknowledgement_List.pdf.

## Notes

### Author Declarations

The BLSA research protocols were conducted in accordance with United States federal policy for the protection of human research subjects contained in Title 45 Part 46 of the Code of Federal Regulations (45 CFR 46), approved by local institutional review boards (IRB), and all participants gave written informed consent at each visit. The BLSA PET substudy is governed by the IRB of the Johns Hopkins Medical Institutions, and the BLSA study is overseen by the National Institute of Environmental Health Sciences IRB. The ADNI study was approved by the IRBs of all of the participating institutions, and all participants gave written informed consent.

### Summary of Updates

10-year cardiovascular disease risk was added as a covariate in all statistical analyses. Reference region used for ADNI florbetapir PET data was a composite region, not whole cerebellum as incorrectly stated in the previous version. ADNI flortaucipir data is now normalized using an inferior cerebellar gray matter reference region (previous version was based on an atlas-based cerebellar cortex reference). Sensitivity analyses with continuous amyloid and alternative reference regions were added.

