## Supplementary Material for "Causal links among amyloid, tau, and neurodegeneration"

### Supplementary Materials

#### Vascular risk variables

We assembled the following variables at or closest to the baseline FTP PET visit included in our analyses: body mass index (BMI), systolic blood pressure (SBP), diastolic blood pressure (DBP), fasting glucose, total cholesterol, high-density lipoprotein (HDL) cholesterol, low-density lipoprotein (LDL) cholesterol, total triglycerides, hypertension diagnosis, diabetes diagnosis, medication use for hypertension, medication use for diabetes, heavy alcohol use (in BLSA) or alcohol abuse (in ADNI), and current smoking.

We used age, sex, total cholesterol, HDL cholesterol, medication use for hypertension, SBP, diabetes diagnosis, and current smoking to compute a 10-year cardiovascular disease (CVD) risk score using the equations reported by D'Agostino et al. based on the Framingham Heart Study.[1] This risk score reflects the probability of developing CVD (i.e., coronary heart disease, cerebrovascular disease, peripheral vascular disease, and heart failure) within 10 years.

*BLSA.* Participants were determined to exhibit heavy alcohol use if they reported drinking > 14 drinks for men and > 7 drinks for women in a typical week over the past 12 months. These thresholds were selected based on the National Institute on Alcohol Abuse and Alcoholism (NIAAA) criteria for heavy alcohol use.[2] If participants reported no drinking in the last 12 months or if their typical weekly consumption was below the thresholds mentioned above, they were considered to not exhibit heavy alcohol use. Otherwise, heavy alcohol use status was considered unknown. Remaining vascular risk variables were assembled and 10-year CVD risk was computed as described previously.[3]

*ADNI.* To determine the presence or absence of hypertension, diabetes, and smoking at each visit, we primarily relied on medical history (binary variable HMHYPERT in MODHACH.csv, binary variable MH16SMOK in MEDHIST.csv, text variable MHCOMMEN in MEDHIST.csv, and text variable MHDESC in RECMHIST.csv) where available. To ascertain hypertension diagnosis, we mined the text variables for the keywords "hypertension", "HTN", "elevated/high blood pressure/BP", allowing for minor spelling errors and variation (e.g., "B/P", "hyoertension"). To ascertain diabetes diagnosis, we mined the text notes for words starting with "diabet" (to encompass "diabetes" and "diabetic"). We considered any match preceded by "no", "not", "not diagnosed", "borderline", "intermittent", "pre", or followed by "borderline", to indicate the absence of a diagnosis. To ascertain smoking status, we mined the text notes for words starting with "smok" (to encompass "smoking" and "smoker") or "cigar" (to encompass "cigar" as well as "cigarette"). If any of the three medical history variables indicated the presence of hypertension or smoking, or either of the two text variables indicated the presence of diabetes, or the participant reported using medication for the condition (as mined from the text variable CMREASON in RECCMEDS.csv), the participant was considered to have the condition at the associated visit. Otherwise, if any of the text variables indicated the absence of a diagnosis (i.e., "borderline hypertension" or "not diagnosed diabetes"), the participant was considered to not have the condition at the associated visit. Visits prior to the absence of hypertension, hypertension medication use, or diabetes were imputed to not have the condition, and visits following a diagnosis of hypertension, hypertension medication use, diabetes, or smoking were imputed to have the condition. If the condition at a given visit could not be ascertained through medical history, medication use, or by longitudinal imputation, we considered the participants to have hypertension at a given visit if they had suprathreshold blood pressure measurements at the visit (SBP ≥ 130 mmHg or DBP ≥ 80 mmHg), to have diabetes if they had a fasting glucose measurement > 125 mg/dL, and to be non-smokers. To obtain blood pressure measurements, we primarily relied on VITALS.csv (continuous variables VSBPSYS and VSBPDIA), but if they were not available in this file, we used the pre- and post-FBP PET scan blood pressure measurements available in AV45VITALS.csv (continuous variables PRESYSTBP, PREDIABP, POSTSYSTBP, and POSTDIABP). Fasting glucose measurements collected prior to FDG PET scans were obtained from PETMETA_ADNI1.csv, PETMETA_ADNIGO2.csv, and PETMETA3.csv (continuous variable PMBLGLUC). Hypertension or diabetes diagnoses determined based on suprathreshold blood pressure and fasting glucose measurements were propagated forward in time within each participant to impute missing values.

Total cholesterol, HDL cholesterol, LDL cholesterol, and total triglycerides were measured using Nightingale Health's Nuclear Magnetic Resonance metabolomics platform[4] (variables TOTAL_C, HDL_C, LDL_C in ADNINIGHTINGALELONG_05_24_21.csv). Cholesterol and triglyceride measurements were converted from mmol/L to mg/dL by multiplying by 38.67 and 88.57, respectively.

Alcohol abuse was determined using the Diagnostic and Statistical Manual of Mental Disorders, fourth edition (DSM-IV)[5] criteria (binary variable MH14ALCH in MEDHIST.csv).

BMI was computed in kg/m$^2$ using height and weight information in VITALS.csv, paying attention to the units (variables VSHEIGHT, VSHTUNIT, VSWEIGHT, VSWTUNIT). We flagged two height measurements in our dataset that were above 85 and whose units were recorded as inches: 156 in and 193 in. We treated these as 156 cm and 193 cm instead.

We imputed any remaining missing values as follows: first, we carried forward in time the latest available value prior to the missing value within each participant. Next, we carried backward in time the first available value preceding the missing value within each participant. Finally, for the purpose of calculating the CVD risk score only, we imputed any remaining missing binary variables as 0 (i.e., absence of condition) and continuous variables at their mean values in the larger cross-sectional ADNI sample (total cholesterol at 192.6 mg/dL, HDL cholesterol at 59.4 mg/dL, and SBP at 135.5 mmHg).

Vascular variables are summarized in Supplementary Table 1. We compared A– and A+ groups within each sample using Wilcoxon rank-sum test for continuous variables and Fisher's exact test for binary variables. Difference between amyloid groups in the prevalence of hypertension was statistically significant in BLSA (P = 0.0054), with the A– group having higher prevalence. Difference between amyloid groups in CVD risk was statistically significant in ADNI ($P = 0.0042$), with the A– group having higher CVD risk. We did not observe any other statistically significant amyloid group differences.

**Supplementary Table 1. Summary of vascular variables.**

| | BLSA | | ADNI | |
|---|---|---|---|---|
| | **A–**, *n* = 71 | **A+**, *n* = 32 | **A–**, *n* = 61 | **A+**, *n* = 61 |
| Systolic blood pressure (mmHg) | 120 (111, 131) | 118 (109, 130) | 135 (127, 145) | 129 (120, 144) |
| Diastolic blood pressure (mmHg) | 66 (60, 73) | 64 (59, 71) | 76 (67, 84) | 72 (69, 80) |
| Total cholesterol (mg/dL) | 182 (164, 210) | 179 (156, 199) | 192 (167, 215)[a] | 186 (160, 210)[a] |
| HDL cholesterol (mg/dL) | 60 (54, 74) | 64 (52, 75) | 57 (48, 66)[a] | 60 (48, 70)[a] |
| Triglycerides (mg/dL) | 88 (70, 118) | 89 (74, 117) | 107 (78, 138)[a] | 81 (72, 116)[a] |
| Body mass index (kg/m$^2$) | 27 (24, 31) | 26 (23, 28) | 27 (25, 30) | 26 (23, 29) |
| Fasting glucose (mg/dL) | 95 (90, 102) | 92 (86, 102) | 93 (85, 106)[b] | 96 (89, 107) |
| Hypertension diagnosis | 44 (62%) | 10 (31%) | 53 (87%) | 47 (77%) |
| Diabetes diagnosis | 9 (13%) | 2 (6%) | 11 (18%) | 10 (16%) |
| Current smoking | 25 (35%) | 10 (31%) | 17 (28%) | 16 (26%) |
| Heavy alcohol use/alcohol abuse[c] | 6 (9%)[d] | 4 (12%) | 3 (6%)[a] | 1 (3%)[a] |
| 10-year cardiovascular disease risk | 0.16 (0.10, 0.23) | 0.13 (0.08, 0.18) | 0.28 (0.17, 0.37) | 0.17 (0.11, 0.28) |

Abbreviations: A– = amyloid negative, A+ = amyloid positive, ADNI = Alzheimer's Disease Neuroimaging Initiative, BLSA = Baltimore Longitudinal Study of Aging.

[a] Lipid measurements and alcohol abuse status were missing for 13 A– and 24 A+ participants in ADNI.

[b] Fasting glucose measurements were missing for 2 A– participants in ADNI.

[c] Alcohol use reflects heavy alcohol use (determined using National Institute on Alcohol Abuse and Alcoholism's thresholds for number of drinks per week) in BLSA and alcohol abuse (determined according to Diagnostic and Statistical Manual of Mental Disorders, fourth edition criteria) in ADNI.

[d] Heavy alcohol use status was missing for 3 A– participants in BLSA.

### Software

We used https://causalfusion.net/[6] to verify the logical expressions for the conditional independencies implied by our DAGs. All statistical analyses were conducted in R (https://cran.r-project.org, version 4.0.3). We used the bnlearn[7] package to test for conditional independence, lmerTest[8] to fit the linear mixed effects models, performance[9] to check model diagnostics, mediation[10] to perform the mediation analyses, tidyverse[11] for data wrangling and plotting, ggExtra[12], ggthemes[13], and wesanderson[14] to generate plots, ggpubr[15] to create panel figures, stargazer[16] to tabulate model results, and knitr[17] to generate the manuscript directly incorporating results from R.

**Causal mediation analysis results in tabular format**

The causal mediation analysis results presented in the main text are summarized below in tabular format. As detailed in the Methods section, these results are based on dichotomous amyloid group determined using a cerebellar gray matter reference region for PiB PET in BLSA and a composite reference region for FBP PET in ADNI. ADE and ACME reflect the direct and indirect effects of amyloid on the outcome, respectively.

**Supplementary Table 2a.** Causal mediation analysis results for the model investigating the relationships among amyloid group (exposure) and tau in the entorhinal cortex (mediator) and the inferior temporal gyrus (outcome) in BLSA and ADNI. Both the mediator and the outcome models were adjusted for age, sex, *APOE* $\varepsilon 4$ positivity, education, and CVD risk. ACME = average causal mediation effect, ADE = average direct effect.

| | **BLSA** | **ADNI** |
|---|---|---|
| **$ACME_{A-}$** | 0.27 ($P = 0.0028$) | 0.12 ($P = 0.0092$) |
| **$ACME_{A+}$** | 0.61 ($P = 0.0028$) | 0.41 ($P = 0.0088$) |
| **$ADE_{A-}$** | 0.30 ($P = 0.038$) | 0.28 ($P = 0.14$) |
| **$ADE_{A+}$** | 0.64 ($P < 0.001$) | 0.57 ($P < 0.001$) |
| **Total effect** | 0.90 ($P < 0.001$) | 0.69 ($P < 0.001$) |

**Supplementary Table 2b.** Causal mediation analysis results for the model investigating the relationships among amyloid group (exposure), tau in the inferior temporal gyrus (mediator), and cerebral blood flow (in BLSA) or glucose metabolism (in ADNI) in the inferior temporal gyrus (outcome). Both the mediator and the outcome models were adjusted for age, sex, *APOE* $\varepsilon 4$ positivity, education, and CVD risk. ACME = average causal mediation effect, ADE = average direct effect.

| | **BLSA** | **ADNI** |
|---|---|---|
| **ACME** | -0.28 ($P = 0.021$) | -0.14 ($P = 0.062$) |
| **ADE** | -0.37 ($P = 0.12$) | 0.080 ($P = 0.70$) |
| **Total effect** | -0.65 ($P = 0.0072$) | -0.059 ($P = 0.76$) |

**Supplementary Table 2c.** Causal mediation analysis results for the model investigating the relationships among amyloid group (exposure), tau in the inferior temporal gyrus (mediator), and inferior temporal gyrus volume (outcome) in BLSA and ADNI. Both the mediator and the outcome models were adjusted for age, sex, *APOE* $\varepsilon 4$ positivity, education, and CVD risk. ACME = average causal mediation effect, ADE = average direct effect.

| | **BLSA** | **ADNI** |
|---|---|---|
| **ACME** | -0.14 ($P = 0.19$) | -0.24 ($P < 0.001$) |
| **ADE** | -0.13 ($P = 0.63$) | -0.29 ($P = 0.15$) |
| **Total effect** | -0.26 ($P = 0.24$) | -0.53 ($P = 0.0084$) |

**Sensitivity of results to dichotomous versus continuous use of amyloid**

We repeated the causal mediation analyses using a continuous amyloid measure (mean cortical PiB DVR computed using a cerebellar gray matter reference region or global cortical FBP SUVR index computed using a composite reference region) instead of binary amyloid status. We denote the mean value of the continuous amyloid measurement in the A– and A+ groups as $\mu_{A-}$ and $\mu_{A+}$. In the first causal mediation analysis involving an interaction between amyloid and EC tau, we computed $ACME_{A-}$ as the expected difference in the outcome while the continuous amyloid measure is fixed at $\mu_{A-}$while the mediator (tau) changes from the value attained or that would have been attained while the continuous amyloid measure is $\mu_{A-}$ to the value under $\mu_{A+}$. We computed $ADE_{A-}$ as the expected difference in the outcome as a result of changing the continuous amyloid variable from $\mu_{A-}$ to $\mu_{A+}$ while keeping the mediator at the value observed or that would have been observed when the amyloid measure is equal to $\mu_{A-}$. $ACME_{A+}$ and $ADE_{A+}$ were defined similarly in this continuous amyloid setting. We note that we could have chosen any two arbitrary operating points for defining ACME and ADE in the continuous amyloid analysis; we decided to use $\mu_{A-}$ and $\mu_{A+}$ to enable the direct comparability of the estimates between the dichotomous and continuous amyloid models.

Results based on continuous amyloid were highly consistent with those based on binary amyloid status (Supplementary Tables 3a–c). We observed the following differences in statistical significance: in the models with continuous amyloid measures, $ADE_{A+}$ for the ITG tau analysis in ADNI was no longer statistically significant ($ADE_{A+}$ = 0.22, $P$ = 0.24). In the BLSA ITG CBF analysis, ACME was no longer statistically significant (ACME = -0.19, $P$ = 0.087) but ADE became statistically significant (ADE = -0.63, $P$ = 0.0036). The total effect for the ITG volume analysis (total effect = -0.38, $P$ = 0.033) also became statistically significant.

Our causal mediation analysis findings were generally robust to dichotomous versus continuous modeling of amyloid, with a few notable differences. In the continuous amyloid model in ADNI, the direct effect of amyloid level on ITG tau among individuals who have levels of EC tau observed among those with elevated amyloid was no longer statistically significant. Given that our ADNI sample consists mostly of individuals with MCI, this finding may suggest that the propagation of tau from EC to ITG is largely facilitated by the presence of amyloid plaques rather than their amount among individuals with MCI, who are already likely to have elevated amyloid levels. On the other hand, using continuous amyloid measures yielded a statistically significant direct instead of indirect effect of amyloid level on ITG CBF in BLSA, suggesting that the level, rather than merely the presence, of amyloid burden might impact proxies of neurodegeneration among cognitively normal individuals.

**Supplementary Table 3a.** Causal mediation analysis results for the model investigating the relationships among continuous amyloid measure (exposure) and tau in the entorhinal cortex (mediator) and the inferior temporal gyrus (outcome) in BLSA and ADNI. Both the mediator and the outcome models were adjusted for age, sex, *APOE* $\varepsilon$4 positivity, education, and CVD risk. ACME = average causal mediation effect, ADE = average direct effect.

| | **BLSA** | **ADNI** |
|---|---|---|
| **$\mathrm{ACME}_{A-}$** | 0.37 ($P = 0.0024$) | 0.29 ($P < 0.001$) |
| **$\mathrm{ACME}_{A+}$** | 0.52 ($P = 0.0028$) | 0.42 ($P < 0.001$) |
| **$\mathrm{ADE}_{A-}$** | 0.27 ($P = 0.059$) | 0.091 ($P = 0.57$) |
| **$\mathrm{ADE}_{A+}$** | 0.43 ($P < 0.001$) | 0.22 ($P = 0.24$) |
| **Total effect** | 0.79 ($P < 0.001$) | 0.51 ($P = 0.0016$) |

**Supplementary Table 3b.** Causal mediation analysis results for the model investigating the relationships among continuous amyloid measure (exposure), tau in the inferior temporal gyrus (mediator), and cerebral blood flow (in BLSA) or glucose metabolism (in ADNI) in the inferior temporal gyrus (outcome). Both the mediator and the outcome models were adjusted for age, sex, *APOE* $\varepsilon$4 positivity, education, and CVD risk. ACME = average causal mediation effect, ADE = average direct effect.

| | **BLSA** | **ADNI** |
|---|---|---|
| **ACME** | -0.19 ($P = 0.087$) | -0.12 ($P = 0.13$) |
| **ADE** | -0.63 ($P = 0.0036$) | -0.15 ($P = 0.26$) |
| **Total effect** | -0.83 ($P < 0.001$) | -0.27 ($P = 0.064$) |

**Supplementary Table 3c.** Causal mediation analysis results for the model investigating the relationships among continuous amyloid measure (exposure), tau in the inferior temporal gyrus (mediator), and inferior temporal gyrus volume (outcome) in BLSA and ADNI. Both the mediator and the outcome models were adjusted for age, sex, *APOE* $\varepsilon$4 positivity, education, and CVD risk. ACME = average causal mediation effect, ADE = average direct effect.

| | **BLSA** | **ADNI** |
|---|---|---|
| **ACME** | -0.085 ($P = 0.38$) | -0.23 ($P = 0.0028$) |
| **ADE** | -0.30 ($P = 0.18$) | -0.27 ($P = 0.087$) |
| **Total effect** | -0.38 ($P = 0.033$) | -0.50 ($P < 0.001$) |

**Sensitivity of results to the reference region used for FBP PET in ADNI**

We repeated our causal mediation analyses in ADNI using amyloid group designations based on a whole cerebellar reference region and the corresponding SUVR cutoff of 1.11. Out of 122 participants, six were classified as A– under the whole cerebellum reference but A+ under the composite reference, and two were classified as A+ under the whole cerebellum reference but A– under the composite reference. There were no differences in statistical significance compared to results obtained with amyloid group designations based on a composite reference region and the corresponding SUVR cutoff of 0.78 (Supplementary Tables 4a–c

In the tables below, ADE and ACME reflect the direct and indirect effects of amyloid on the outcome, respectively.

**Supplementary Table 4a.** Causal mediation analysis results for the model investigating the relationships among amyloid group based on a cerebellar reference region (exposure) and tau in the entorhinal cortex (mediator) and the inferior temporal gyrus (outcome) in ADNI. Both the mediator and the outcome models were adjusted for age, sex, *APOE ε*4 positivity, education, and CVD risk. ACME = average causal mediation effect, ADE = average direct effect.

| | **ADNI** |
|---|---|
| **$ACME_{A-}$** | 0.14 ($P = 0.0024$) |
| **$ACME_{A+}$** | 0.45 ($P = 0.0024$) |
| **$ADE_{A-}$** | 0.32 ($P = 0.071$) |
| **$ADE_{A+}$** | 0.63 ($P < 0.001$) |
| **Total effect** | 0.77 ($P < 0.001$) |

**Supplementary Table 4b.** Causal mediation analysis results for the model investigating the relationships among amyloid group based on a cerebellar reference region (exposure), tau in the inferior temporal gyrus (mediator), and glucose metabolism in the inferior temporal gyrus (outcome) in ADNI. Both the mediator and the outcome models were adjusted for age, sex, *APOE ε*4 positivity, education, and CVD risk. ACME = average causal mediation effect, ADE = average direct effect.

| | **ADNI** |
|---|---|
| **ACME** | -0.16 ($P = 0.073$) |
| **ADE** | 0.050 ($P = 0.82$) |
| **Total effect** | -0.11 ($P = 0.58$) |

**Supplementary Table 4c.** Causal mediation analysis results for the model investigating the relationships among amyloid group based on a cerebellar reference region (exposure), tau in the inferior temporal gyrus (mediator), and inferior temporal gyrus volume (outcome) in ADNI. Both the mediator and the outcome models were adjusted for age, sex, *APOE ε*4 positivity, education, and CVD risk. ACME = average causal mediation effect, ADE= average direct effect.

| | **ADNI** |
|---|---|
| **ACME** | -0.26 ($P < 0.001$) |
| **ADE** | -0.36 ($P = 0.073$) |
| **Total effect** | -0.63 ($P = 0.0036$) |

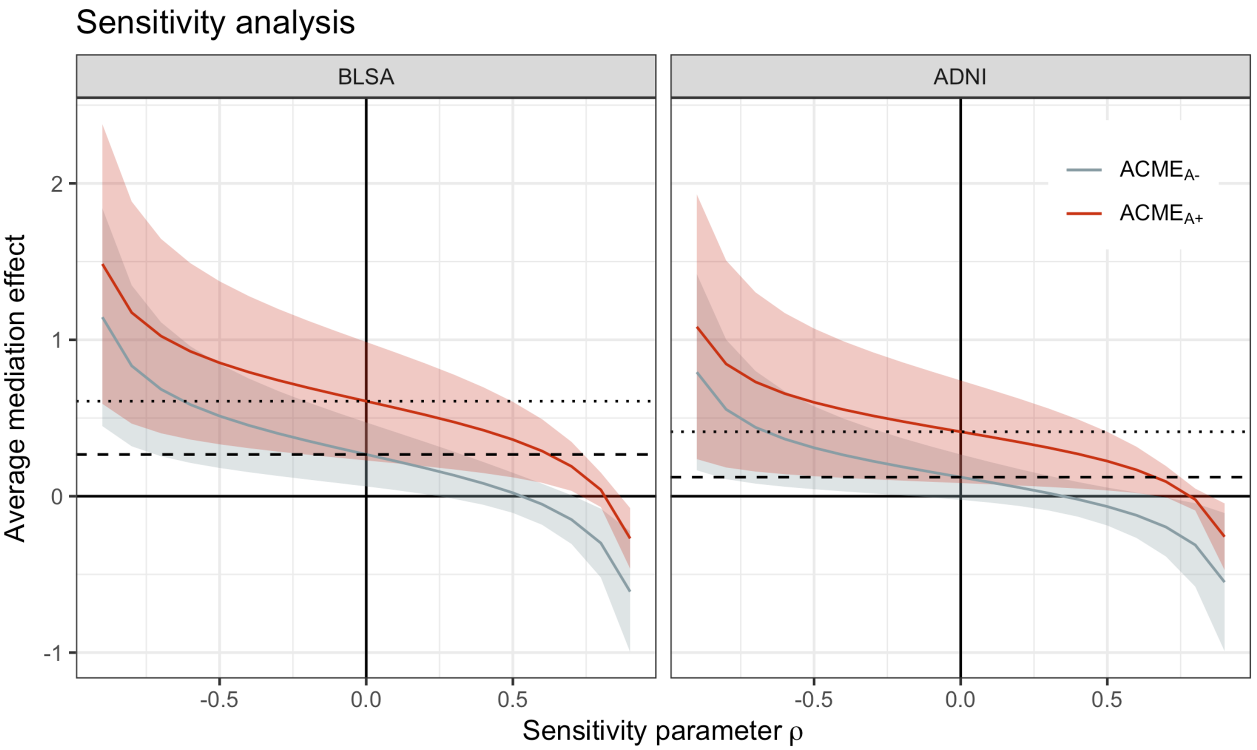


**Supplementary Figure 1. Sequential ignorability assumption sensitivity analyses for the causal mediation models investigating entorhinal cortex tau as a mediator of the association between amyloid group and inferior temporal gyrus tau.** Sensitivity parameter $\rho$ is the simulated correlation between the mediator and the outcome model residuals. The 95% confidence interval of ACME$_{A+}$ does not include the value zero in either data set when $\rho < 0.5$. This indicates that even in the presence of an unspecified confounder accounting for residual correlations up to 0.5 between the mediator and the outcome, our conclusion that ACME$_{A+}$ is statistically significant would hold. ACME = average causal mediation effect, ADNI = Alzheimer's Disease Neuroimaging Initiative, BLSA = Baltimore Longitudinal Study of Aging.

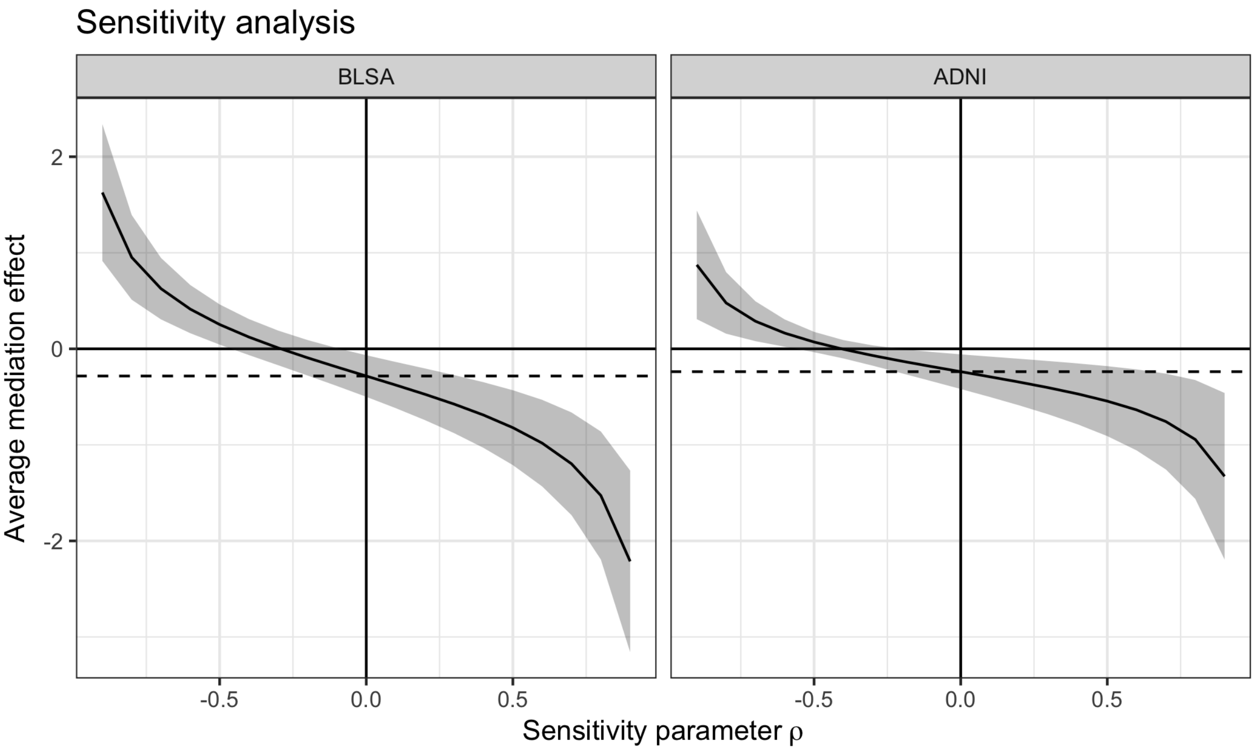


**Supplementary Figure 2. Sequential ignorability assumption sensitivity analysis for the causal mediation models investigating inferior temporal gyrus tau as a mediator of the association between amyloid group and inferior temporal gyrus cerebral blood flow in BLSA (left) or volume in ADNI (right).** Sensitivity parameter $\rho$ is the simulated correlation between the mediator and the outcome model residuals. The 95% confidence interval of ACME includes the value zero in both data sets for values of $\rho$ not too far from zero, indicating the presence of unspecified confounders might yield an ACME that is statistically non-significant. ACME = average causal mediation effect, ADNI = Alzheimer's Disease Neuroimaging Initiative, BLSA = Baltimore Longitudinal Study of Aging.

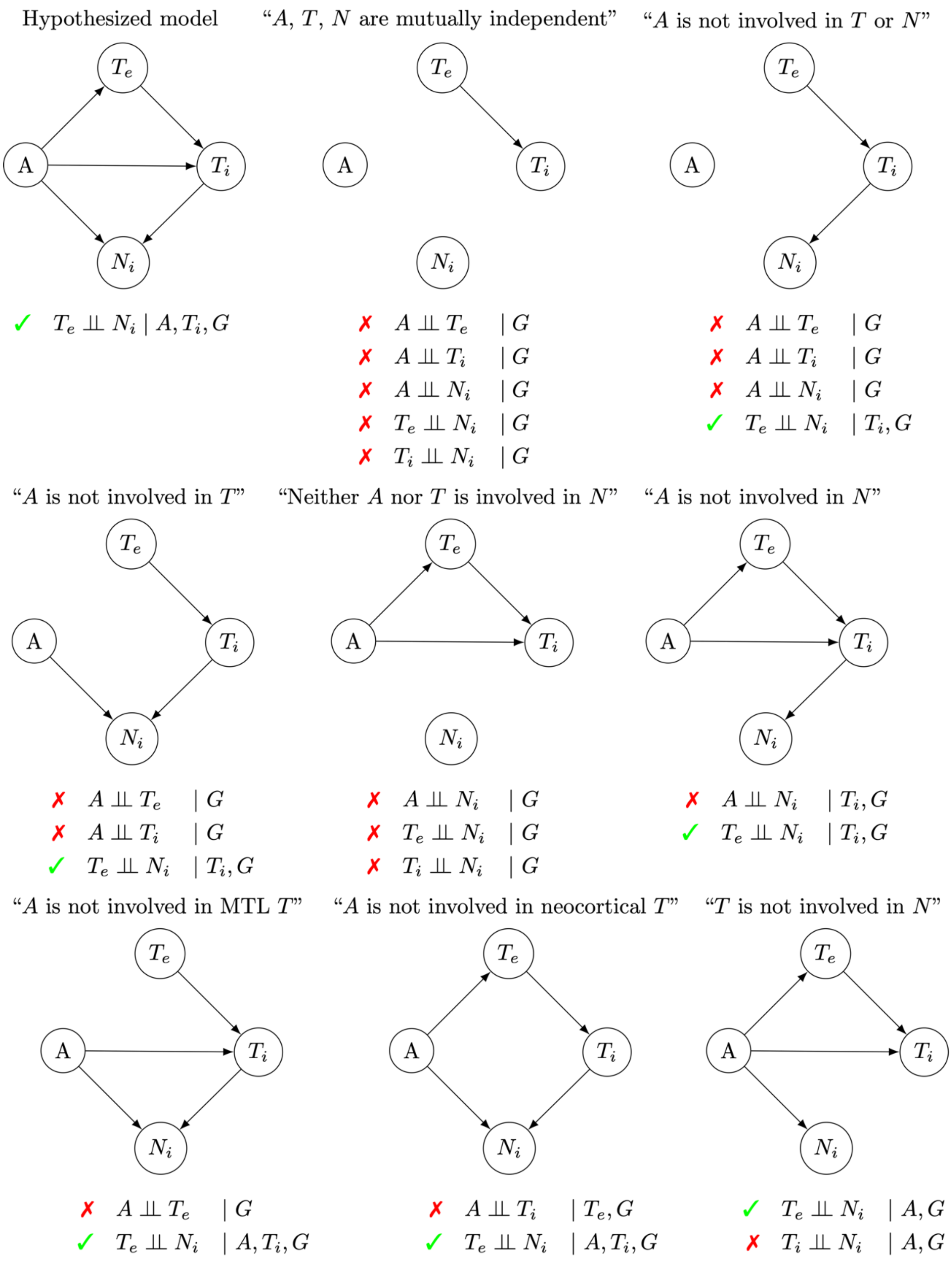


**Supplementary Figure 3. Conditional independencies for more parsimonious directed acyclic graphs (DAGs).** *G* indicates the collection of covariates (age, sex, education, 10-year cardiovascular disease risk, and *APOE* ε4 status). Each of these covariates is a node with an arrow towards each of the four nodes shown in this diagram (nodes for covariates are not shown). The conditional independencies implied by each DAG are listed below the graphs.

Green check mark indicates that the partial correlation is not statistically significant in the observed data (and therefore, the DAG is not inconsistent with the data). Red cross mark indicates that the partial correlation is statistically significant (and therefore, the DAG is inconsistent with the data). $A$ = binary amyloid status, $T_e$ = entorhinal cortex tau, $T_i$ = inferior temporal gyrus tau, $N_i$ = inferior temporal gyrus neurodegeneration.
